# Structure-function coupling reveals seizure onset connectivity patterns

**DOI:** 10.1101/2022.09.21.22280190

**Authors:** Christina Maher, Arkiev D’Souza, Michael Barnett, Omid Kavehei, Chenyu Wang, Armin Nikpour

## Abstract

The implications of combining structural and functional connectivity to quantify the most active brain regions in seizure onset remain unclear. We obtained structural connectomes from diffusion MRI (dMRI) and functional connectomes from electroencephalography (EEG) to assess whether high structure-function coupling corresponded with the seizure onset region. We mapped individual electrodes to their nearest cortical region to allow for a one-to-one comparison between the structural and functional connectomes. A seizure laterality score and expected onset zone were defined. The patients with well-lateralised seizures revealed high structure-function coupling consistent with the seizure onset zone. However, a lower seizure lateralisation score translated to reduced alignment between the high structure-function coupling regions and the seizure onset zone. This feasibility study tested a new model for incorporating dMRI in clinical practice. We illustrate that dMRI, in combination with EEG, can improve the identification of the seizure onset zone. Our model may be valuable in enhancing ultra-long-term monitoring by indicating optimal, individualised electrode placement.

## 1. Introduction

The assessment of patients with focal epilepsy using a combination of structural data derived from diffusion MRI (dMRI) and functional data from electroencephalography (EEG) is gaining increased appeal [1, 2, 3]. In the brain, structural connectivity refers to an anatomical link between two or more brain regions. Connnectomes generated from diffusion MRI, can represent the strength of structural connectivity between specific brain regions. Functional connectivity is inferred from the spatio-temporal relationship between electrophysiological signals from two or more structurally discrete regions [4]. Structural connectivity is believed to give rise to functional and network behaviour [5]. In a mechanistic sense, the composition of white matter can be expected to influence the flow of activity and connectivity between neuronal populations. Therefore, if EEG functions as a tool to observe the flow of activity, the connectivity measurements from EEG can be presumed to closely resemble connectivity measurements from structural MRI. In epilepsy, structure-function coupling is proposed to have a role in identifying seizure propagation patterns [6, 7], seizure generalisation [1] and predicting post-surgery seizure freedom [8, 9]. Diffusion MRI (dMRI) derived tractography enables the quantification of structural connectivity between different brain regions. However, in epilepsy, dMRI is held to be in the experimental realm [2]. Though there is consensus on the significance of structural connectivity information in patient diagnosis, the utility of dMRI as a routine clinical test has not been realised. Additional research is needed to investigate the value of dMRI in combination with routinely collected data such as EEG. Further, the feasibility of user-friendly tools for deploying dMRI pipelines must be assessed.

Several works employ functional MRI (fMRI) to represent functional connectivity [10, 11, 12] alongside structural connectivity from dMRI. However, given EEG is routinely collected in epilepsy clinics, it may be a more accessible and practical alternative to fMRI, which has inherent, poor temporal resolution relative to EEG. White matter connectivity and information flow between specific brain regions has been linked to scalp EEG characteristics in healthy populations [13, 14]. Further, EEG has been used to produce an individualised connectivity fingerprint that is robust across recordings [15], rendering its utility as a patient-specific, analytical network measure that can address the heterogeneous nature of focal epilepsy.

Discerning the seizure onset pattern and epileptogenic zone has been shown to improve the prognosis of post-surgical outcomes [16], and EEG and dMRI can aid this goal. A study on the role of scalp EEG in predicting post-surgical seizure outcomes showed abnormal MRI was valuable in ambiguous cases containing bilateral interictal epileptiform discharges [17], suggesting MRI may enhance prediction of seizure freedom. In another study of seven patients being evaluated for epilepsy, lesional and non-lesional MRIs were combined with high and low frequency bands from high density EEG (HDEEG) [18]. The authors showed that the absence of structural support was related to significantly reduced functional connectivity in high frequency bands. High frequency oscillations observed on scalp EEG are increasingly recognised as a hallmark of lesional epilepsy [19]. These works highlight the advantages of combining dMRI with EEG to detect aberrations that typically may only be partly revealed by one modality.

The majority of works that blend multimodal information from dMRI and EEG focus on source localisation techniques [20, 21, 22], using a digitiser to map electrodes coordinates to the scalp which can be time-consuming. Others produced an automated, individualised localisation tool to map electrodes from high density EEG (HDEEG) to the scalp only, without extending the mapping to the cortex [23]. Many prior works favour the combination of stereo EEG with dMRI [6, 9, 24, 25], or only explored normal (non-ictal) awake EEG data with dMRI [26].

Several methods for electrical source localisation, which utilise a range of forward and inverse solutions, have been proposed and evaluated [27, 28, 29]. The current study is distinguished from those prior works for the following reasons. In a larger cohort, we sought to apply our model [30], which maps cortex regions to individual electrodes. We aimed to understand whether a patient-specific, structure-function coupling pattern could be observed without requiring manual digitisation of electrodes or applying one of the several forward and inverse solutions. We specifically examined the seizure onset period (regardless of wakefulness state). We aimed to validate the feasibility of our model as a clinically translatable method to leverage the potential of dMRI, with the view of elevating it to the established state currently held by structural MRI (i.e. T1) [31]. The dMRI component of our tool was designed to be deployed on a clinician’s computer, allowing straightforward data processing from new patients (with ethics approval).

The contribution of this work is twofold: 1. We extend the application of our spatial mapping model to a new patient cohort, highlighting consistent between-patient variance in region to electrode mapping 2. We add to the growing body of research showing that connectivity data derived from structural MRI may augment scalp EEG observations for certain patients; acting as an additional tool during the diagnosis stage.

## 2. Materials and Methods

### 2.1. Participants and Data

Nine adults with focal epilepsy were recruited from the Comprehensive Epilepsy Centre at the Royal Prince Alfred Hospital (RPAH, Sydney, Australia), and MRI was performed at the Brain and Mind Centre (Sydney, Australia). Inclusion criteria were adults diagnosed with non-lesional focal epilepsy, aged 18-60, presenting without surgery, with a minimum of two recorded seizures, and who were willing and able to comply with the study procedures for the duration of their participation. Exclusion criteria were pregnant women and individuals with intellectual disabilities. Ethical approval was obtained from the RPAH Local Health District (RPAH-LHD) ethics committee (see Institutional Review statement in section 5). A schematic of the data processing steps is shown in Fig. 1.

**Figure 1.**
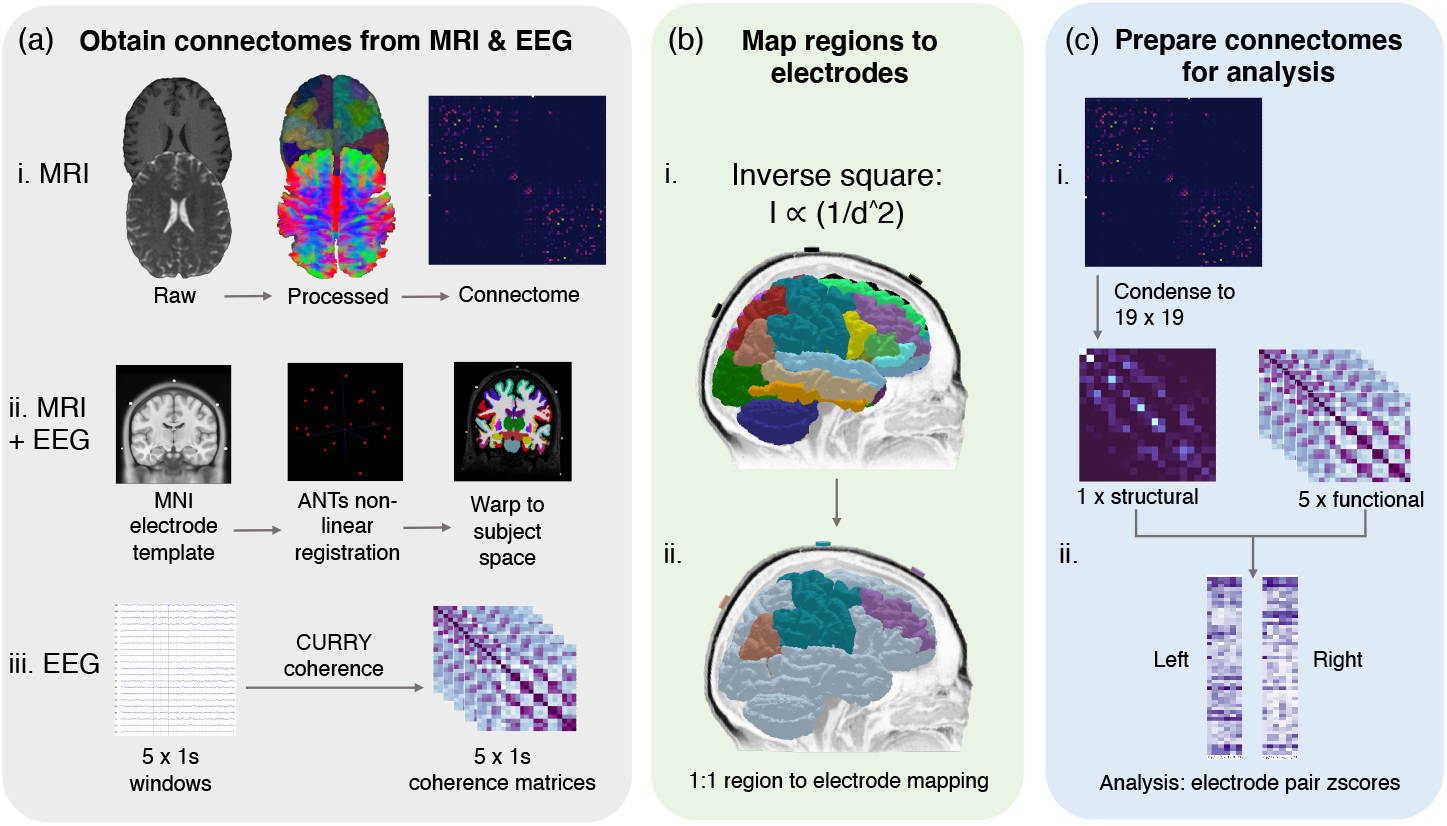
Schematic of data processing to obtain structural and functional connectomes. To obtain the structural connectomes (a, i), the dMRI was processed, and anatomically-constrained probabilistic tractography was conducted as outlined in Section 2.3. Structural regions of interest were based on the DK atlas. To obtain the automated, subject-specific electrode warp (a, ii), electrodes were first mapped to the standard MNI template in Curry. Next, ANTs non-linear registration was used to warp the electrodes from the MNI template space to the subject space. To obtain the functional connectomes (a, iii), the first 5 seconds of a given seizure were selected using one-second windows. Each one-second window was processed using Curry’s sensor coherence algorithm, producing a 21×21 coherence matrix (the reference electrodes were then removed before statistical analysis). To map each cortical region to the nearest electrode, we applied our inverse square script (b, i) to each subject’s electrode warp file and their Freesurfer-derived parcellation file (subcortical regions were removed during this process). The result was a corresponding electrode to each of the 70 regions, i.e. F7/L.LOFG (b, ii.). Next, the 70×70 region matrix was condensed to 19×19 (c, i) to match the dimensions of the EEG functional matrix. Specifically, the values of all regions corresponding to a given electrode pair were summed, and the final value was used as the connectivity value for that electrode pair in the structural matrix. Lastly, Z-scores were computed for all connectivity values in the structural and functional matrices.

### 2.2. Image acquisition

Image acquisition was described previously [32]. Briefly, all scans were acquired on the same GE Discovery™ MR750 3T scanner (GE Medical Systems, Milwaukee, WI). The following sequences were acquired for each participant: Pre-contrast 3D high-resolution T1-weighted image (0.7mm isotropic) using fast spoiled gradient echo (SPGR) with magnetisation-prepared inversion recovery pulse (TE/TI/TR=2.8/450/7.1ms, flip angle=12); and axial diffusion-weighted imaging (2mm isotropic, TE/TR=85/8325ms) with a uniform gradient loading (*b*=1000s/mm^2^) in 64 directions and 2 *b* 0s. An additional *b*0 image with reversed phase-encoding was also acquired for distortion correction [33].

### 2.3. Image processing to obtain structural connectomes

The T1 images were processed using a modified version of Freesurfer’s recon-all (v6.0) [34], alongside an in-house skull-stripping tool (Sydney Neuroimaging Analysis Centre). Each subject was inspected, and minor segmentation errors were manually corrected. A 5 tissue-type (5TT) image [35] was generated using MRtrix3 [36]. The T1 image was registered to the mean *b*0 image; the warp was used to register the 5TT image, and the Desikan-Killiany (DK) parcellation image to the diffusion image.

Diffusion image processing was conducted using MRtrix3 [36]. The diffusion pre-processing included motion and distortion correction [33, 37], bias correction using ANTs [38]. The *dhollander* algorithm [39] was used to estimate the response functions of the white matter, grey matter, and cerebral spinal fluid, from which constrained spherical deconvolution was used to estimate the fibre orientation distributions using MRtrix3Tissue [36]. The intensity of the white matter fibre orientation distributions was normalised [36], and used for anatomically constrained whole-brain tractography [40] (along with the registered 5TT image). The tractography specifications were as follows: 15 million tracks were generated, iFOD2 probabilistic fibre tracking [41], dynamic seeding [42], maximum length 300 mm, backtrack selected and crop at grey-matter-white-matter interface selected. For quantitative analysis, the corresponding weight for each streamline in the tractogram was derived using SIFT2 [42]. The streamlines and corresponding SIFT2 weights were used to create a weighted, undirected structural connectome using the registered parcellation image.

### 2.4. EEG acquisition

The EEG recordings were derived from ward recordings conducted during the patients’ stay at the RPAH. The EEG was recorded using Compumedics hardware and software. The ward nurse applied the individual electrodes to the patient’s head in the standard 10/20 format using the gold standard measurement process. Once the routine clinical recording was complete, the raw EEG files were obtained, and the seizure segments annotated by the EEG technician and reviewed by the neurologist. All seizures were then analysed in Curry to obtain the functional connectomes.

### 2.5. EEG processing to obtain functional connectomes

CURRY 8 software (Compumedics Neuroscan) was used to pre-process the EEG and obtain the sensor-based coherence matrices used to represent functional connectomes. First, we applied Curry’s automated artifact reduction and filtering tool to obtain a clean signal, followed by sensor-based coherence maps. Specifically, using one-second non-overlapping windows starting from the annotated seizure onset time, we obtained the sensor-based coherence maps for the first five seconds of each seizure. The coherence maps were 21*×*21 matrices; row and column headers represented single electrodes shown in Fig. 1 (a, iii).

Per the Curry 8 manual, the coherence calculation is as follows: “channel signals are zero-padded, fast Fourier transformed (FFT), correlations are computed, then back transformed, normalised and searched for the lags in of the maximum overlap”. Following the FFT step, the coherence maps were computed from the cross-spectral densities **Gxy** and auto-spectral densities **Gxx** and **Gyy** of the channels x and y, using the equation **Cxy** = (**Gxy** * **Gxy**)/(**Gxx** * **Gyy**).

Each electrode pair’s corresponding value was a composite of the normalised maximum similarity between the waveforms and the time-shift (delay) when the maximum similarity occurred. The electrode pair value represented the highest percentage of coherence achieved by that electrode pair in the one-second window after factoring in the signal time lag between the two electrodes.

### 2.6. Mapping cortical structures to the nearest electrode

We used our method [30] to produce an electrode warp in the 10/20 format, and to map each electrode to the nearest cortical structure in subject space. Briefly, a nonlinear registration was applied to each participant to warp the 21 electrodes from the standard MNI template space to each participant’s T1 image (that had been registered to the diffusion image space). Cortical regions were labelled according to the DK atlas [43] (using FreeSurfer [34]) to match the image processing. We used our inverse square (IS) method, which was previously shown to produce the least between-subject variance in the region to electrode mapping. Our method incorporates the inverse square formula, shown in Fig. 1 (b, i), which holds that the surface area of a sphere increases with the square of the radius. Thus the inverse square method enabled the inclusion of the voxel intensity in the MRI. Voxel intensity can be used to represent the topological arrangement of the cortex and supports postulation of the strength of an EEG signal from a given region relative to the region’s distance from the scalp. Thus, in combination with the subject-specific electrode warp, we incorporated the inverse square method to produce a subject-specific, one-to-one mapping of each cortical region to its nearest electrode.

### 2.7. Mapping the structural connectome to the functional connectome

The 84*×*84 matrix of regions used as the structural connectome was derived from the 84 node DK atlas. To allow for one-to-one comparisons between the structural (84×84) and functional connectomes (21*×*21), we computed a subject-specific condensed structural connectome as follows: for each region in each subject, the inverse square method determined the nearest electrode. We took all regions nearest to a given electrode pair (i.e. FP1 and Fz) and summed the structural connectome values of those regions (from one half of the matrix). Subcortical regions were not assigned electrodes as their physical distance from the scalp and positioning below other cortical regions deemed them inaccessible for accurate measurement, and they were removed during the electrode mapping process. The resulting value was used as the cell value for that electrode pair in the condensed version of the structural connectome, which was 19×19. We then removed the diagonal line from the matrix (self correlations) of both the structural and functional connectomes and converted the matrices of both connectome types to a 1D array for statistical analysis.

### 2.8. Statistical analysis of structure-function coupling

To test whether the laterality of the strong connections on both the EEG and MRI matched the patient’s diagnosis, we first split the electrode pairs into left and right hemispheres and removed any cross-hemisphere electrode pairs. For example, if an electrode pair contained two electrodes in the left hemisphere (i.e. FP1-F3) or one left hemisphere and one central electrode (FP1-Fz), the electrode pair was kept. All electrode pairs that crossed from one hemisphere to the other (i.e. F3-F4) were removed. To test whether the highly coherent EEG electrode pairs were congruent with highly connected MRI electrode pairs, we first computed the z-scores for all electrode pairs from both data types. The following z-score arrays were produced: a. 1*×*1D array per hemisphere for the structural connectome and b. 5*×*1D arrays (for each 1 second time window) per seizure, per hemisphere for the functional connectomes.

The neurologist provided a “laterality” score for each patient based on whether the frequently observed seizure onset zone was consistently restricted to one hemisphere. A laterality score of zero represented poorly lateralised seizures, whilst highly lateralised seizures received a score of three. If overall, the patient had late-lateralising seizures, they were classified as being non-lateralised at onset (i.e. a score of 0-1). Each patient was also assigned an “expected onset zone”, predicated on the most frequently observed onset zone observed in all of a patient’s recorded seizures, including seizures that were poorly or non-lateralised.

To preserve only the most robust connections to represent high coherence between two electrodes, a z-score threshold of two (i.e. two standard deviations from the mean) was chosen for the structural connectome, and a threshold of 1.8 was chosen for the functional connectome. Next, the z-scores from the structural and functional connectomes were compared per one-second window from each seizure. If the same electrode pair from the structural and functional connectome contained a z-score between 1.8 and 2, that electrode pair was classified as showing high structure-function coupling (termed “coupled electrode pairs”). Lastly, the neurologist reviewed the raw EEG to confirm whether the electrode pair with high structure-function coupling was congruent with the expected seizure onset zone. All z-scores and statistical analyses were produced in SPSS v28.

## 3. Results

### 3.2. Demographics

Nine patients (6F, mean age 38.8 *±*11.28) were included in this study after meeting the inclusion criteria. The patient characteristics, including seizure onset zone, are shown in Table 1. Three of the nine patients presented with highly lateralised seizures; the other six had a mixture of highly lateralised and poorly lateralised seizures. All patients were diagnosed with focal epilepsy; two had experienced frequent focal to bilateral tonic-clonic (FBTC) seizures, whilst another three had infrequent FBTC seizures (experienced more than one year prior to the EEG recording).

**Table 1.** Characteristics of patients

| Patient | Sex | Classification | MRI diagnosis | Age at MRI | Duration | Drug res. | Handedness |
| --- | --- | --- | --- | --- | --- | --- | --- |
| 1 | F | Left fronto-temporal | Normal | 50-59 | 4 | Y | R |
| 2 | M | Left fronto-temporal | Normal <sup>†</sup> | 40-49 | 28 | Y | L |
| 3 | F | Right frontal | Normal | 40-49 | 10 | N | R |
| 4 | F | Right temporal | Normal | 20-29 | 13 | Y | U |
| 5 | M | Left fronto-temporal | Normal | 30-39 | 15 | Y | R |
| 6 | F | Left occipital | Normal | 40-49 | 35 | Y | R |
| 7 | F | Left fronto-temporal | Normal | 40-49 | 13 | N | R |
| 8 | M | Right fronto-temporal | Normal | 30-39 | 18 | Y | R |
| 9 | M | Right temporal | Normal <sup>‡</sup> | 20-29 | 7 | Y | R |

Key: L: left, R: right, U: unknown

### 3.3. Electrode-region mapping

The regions that displayed the most variance in electrode mapping across all nine patients are listed in Table 2 according to the Freesurfer region names. The majority of the variance appeared to be in the temporal regions. Manual inspection of the warped electrodes on each patient’s scalp, which were overlaid on the cortex regions, indicated that individual scalp and cortex morphology contributed to the model’s determination of the nearest electrode for a given region.

**Table 2.** Between patient region variance in the region to electrode mapping

| Subject no. | 1 | 2 | 3 | 4 | 5 | 6 | 7 | 8 | 9 |
| --- | --- | --- | --- | --- | --- | --- | --- | --- | --- |
| Region name |  |  |  |  |  |  |  |  |  |
| L.rostralanteriorcingulate | FP1 | FP1 | FP1 | FZ | F3 | FP1 | FZ | FP1 | FP1 |
| R.rostralanteriorcingulate | FP2 | FP2 | FP2 | FZ | FP2 | FP2 | FZ | F4 | FP2 |
| L.parsopercularis | F3 | F7 | F7 | F3 | F7 | F7 | T3 | F7 | F3 |
| R.parsopercularis | F8 | F8 | F8 | F8 | F8 | F8 | F4 | T4 | F4 |
| L.insula | T3 | F7 | T3 | F7 | T3 | T3 | T3 | T3 | T3 |
| R.insula | T4 | F8 | F8 | F8 | F8 | T4 | C4 | T4 | T4 |
| L.inferiortemporal | T5 | T5 | T5 | T5 | T3 | T5 | T5 | T3 | T5 |
| R.inferiortemporal | T6 | T4 | T6 | T6 | T4 | T6 | T6 | T4 | T6 |
| L.lateralorbitofrontal | FP1 | F7 | F7 | F7 | F7 | F7 | F7 | F7 | F7 |
| R.lateralorbitofrontal | FP2 | F8 | F8 | F8 | F8 | F8 | F4 | F8 | FP2 |
| L.cuneus | O1 | O1 | O1 | O1 | O1 | O1 | PZ | O1 | O1 |
| R.cuneus | O2 | O2 | O2 | PZ | O2 | O2 | PZ | O2 | O2 |
| L.transversetemporal | T3 | T3 | T3 | C3 | T3 | T3 | T3 | T3 | T3 |
| R.transversetemporal | T4 | T4 | T4 | C4 | T4 | T4 | T4 | T4 | T4 |
| L.caudalanteriorcingulate | FZ | FZ | FZ | FZ | F3 | FZ | FZ | F3 | FZ |
| R.caudalanteriorcingulate | FZ | FZ | FZ | FZ | FZ | FZ | FZ | F4 | FZ |
| L.isthmuscingulate | PZ | PZ | PZ | CZ | PZ | PZ | PZ | PZ | PZ |
| R.isthmuscingulate | PZ | PZ | PZ | CZ | PZ | PZ | PZ | PZ | PZ |
| R.bankssts | T6 | T6 | T4 | T6 | T4 | T6 | T6 | T6 | T6 |
| R.superiorfrontal | FZ | FZ | FZ | FZ | FZ | FZ | FZ | CZ | FZ |
| R.caudalmiddlefrontal | C4 | C4 | C4 | F4 | C4 | C4 | C4 | C4 | C4 |
| R.temporalpole | F8 | F8 | F8 | F8 | F8 | F8 | F8 | T4 | F8 |
| L.supramarginal | C3 | P3 | C3 | C3 | C3 | C3 | C3 | C3 | C3 |
| L.superiorparietal | P3 | PZ | PZ | PZ | PZ | PZ | PZ | PZ | PZ |

Key: L: left, R: right, bankssts: banks of the superior temporal sulcus

### 3.4. Structure-function coupling

The structure-function coupling observed in the nine patients revealed three distinct groups with the following features. The first group (Patients 1-3, Fig 2, a) had the highest laterality scores (L= 3), with coupled electrode pairs that consistently overlapped with the seizure onset zone. The second group (Patients 4-6, Fig 2, b) had less well-lateralised seizures (L= 2 *−* 3), and the coupled electrode pairs overlapped with the onset side but not the exact zone. In Patient 4, only two out of three seizures were highly lateralised (L= 3) while the third was not (L= 2), and the electrode pair (PZ-O2) that did not overlap with the exact seizure onset zone was observed on the poorly lateralised seizure. Patient 4 also had three single electrodes from highly connected MRI and EEG pairs that overlapped inside the seizure onset zone (shown in Appendix A1, b). Patients 5 and 6 were less well lateralised, and highly coupled electrode pairs were present both within and outside the seizure onset zone. In Patient 6, the electrode pair T3-C3 overlapped with the expected onset zone of the one poorly lateralised seizure. The third group (Patients 7-9, Fig 2, c) were considered non-lateralised for most of their seizures. These patients generally displayed highly coupled electrode pairs that were inconsistent with the expected onset zone or had only a single overlapping MRI and EEG electrode rather than a pair (Patient 7). Figure 2 contains a condensed interpretation of the results shown in Appendix A1. The overlapping electrode pairs with MRI z-scores (>2) and EEG z-scores (>1.8) are displayed. The most common seizure onset zone for each patient is also displayed. The detailed results for each patient can be viewed in the supplementary material in Appendix A1.

**Figure 2.**
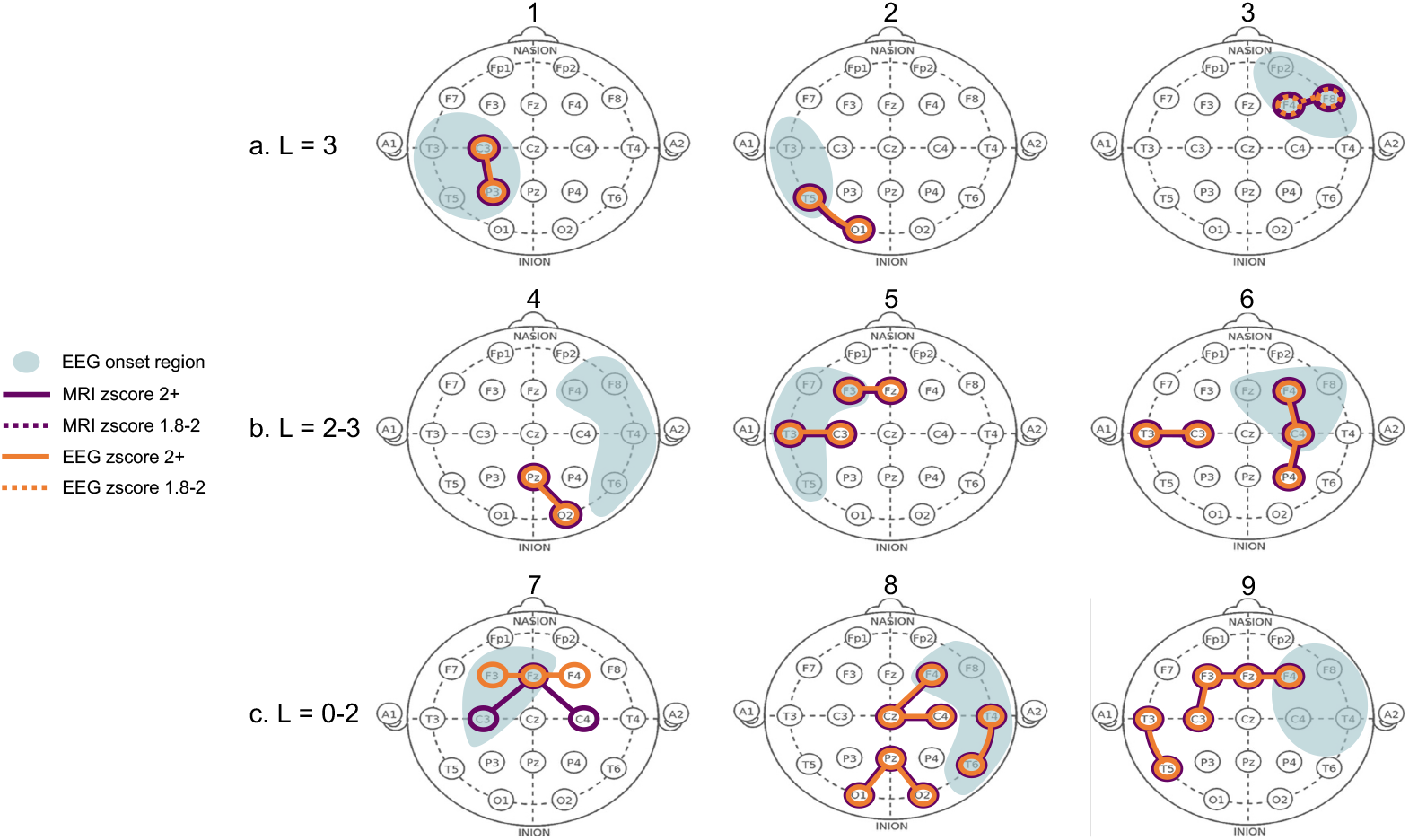
Highly connected electrode pairs in structural and functional connectome. Each “head” shows a schematic of the electrode pairs for each of the nine patients (numbered 1-9 above each head). The “L” value represents each patient’s overall seizure lateralisation score based on their available recorded seizures. A score of zero represented poorly lateralised seizures, whilst highly lateralised seizures received a score of three. The patients’ seizures stratified them into three categories: patients 1-3 had high laterality, patients 4-6 had some seizures that were well lateralised, while others were not, and patients 7-9 had poor laterality in all seizures. The purple lines (and circled electrodes) represent the electrode pairs that displayed strong connectivity (z-scores > 2) in the structural (MRI) connectome. The orange lines (and circled electrodes) represent the electrode pairs that displayed strong connectivity (z-scores > 2) in the functional (EEG) connectome. Dotted lines in either colour represent a z-score of 1.8-2. The blue shading represents the most frequently observed seizure onset zone for a given patient, as observed from their ward EEG recordings. If a seizure did not have a specific onset region within the first 5 seconds, it was considered non-lateralised, even if it displayed late-lateralisation. Purple and orange circled electrodes in the blue shaded areas represent high structure-function coupling in the seizure onset zone.

## 4. Discussion

In this study, we obtained structural and functional connectomes from nine patients to investigate whether our model could uncover the structure-function coupling during seizure onset. We also examined the pattern and congruence of the structure-function coupling with the expected seizure onset zone. The first key finding was that patients with well-lateralised seizures displayed high structural-functional congruence consistent with the expected seizure onset zone. The second key finding was that patients who were not well-lateralised had varying coupled electrodes that were not consistently in the onset zone.

The results indicate that for well-lateralised patients, connectivity data derived from dMRI can be a valuable tool to augment routine EEG observations. However, the dMRI should be interpreted in the context of other routinely collected data from the patient.

Our findings offer some compelling evidence for the use of dMRI in clinical practice. Firstly, in patients with high structure-function coupling in the expected onset zone, dMRI may provide additional support to the EEG observations. A recent work used intracranial EEG (iEEG) and dMRI to explore the relation between structure-function coupling and post-surgery seizure freedom [9]. The authors showed that patients who achieved post-surgery seizure freedom had higher structure-function coupling pre-surgery. However, access to iEEG may not be feasible in the initial diagnosis stage. Additionally, the diagnostic yield of low-density scalp EEG (25 electrodes) has been suggested to be comparable to high-density EEG (256 electrodes) [44]. Taken together, our findings suggest that our model can be used during the diagnosis stage to determine the suitability of surgery for newly-diagnosed patients. Notably, we provide evidence that high structure-function coupling was present for patients regardless of whether they had previously experienced an FBTC seizure. Our finding suggests that a history of infrequent FBTC seizures (present in Patients 1 and 3) does not preclude the patient from having well-lateralised structure-function coupling.

Secondly, in the patients with poorly lateralised seizures, the structure-function coupling was more predominant in the ipsilateral hemisphere. However, the presence of structure-function coupling in the contralateral hemisphere was unsurprising, given their laterality score. In these patients, dMRI may provide additional information that can guide the placement of additional electrodes in longer ward recordings. However, a more extensive structure-function coupling model may be needed to understand whether the poor laterality can be attributed to the equally high structural connectivity in both hemispheres or some other biophysical phenomenon. Further, our model demonstrated a specific region with high structure-function coupling for Patient 8, who was initially considered poorly lateralised on EEG. Such cases highlight that dMRI may offer endorsement of a specific onset zone to support an otherwise inconclusive EEG recording.

Interestingly, the presence of high structure-function coupling in an electrode pair containing a middle electrode (FZ, CZ, PZ) was observed in several patients with a seizure laterality score of 2 or lower. For example, patients 5, 7, 8 and 9 had at least one middle electrode in the electrode pairs that showed high structure-function coupling. Their poor seizure lateralisation could be due to the electrophysiological activity beneath the middle electrode, which may drive the contralateral seizure propagation.

Given the scope of the current work, some methodological considerations may aid the interpretation of the results. Firstly, the mapping method did not appear to impact the results as well-lateralised patients had a similar number of variances in the electrode-to-region mapping as the less well-lateralised patients, for whom the structural data provided little additional information. However, laterality alone may not account for the results of patients who displayed high structure-function coupling congruent with the expected onset zone. Our selection of time window and bandwidth may have impacted the coherence score. The one-second window was perhaps not brief enough to capture the highly coherent initial EEG activity. We observed high coherence in the ipsilateral hemisphere at the 0-100 millisecond (ms) scale for some patients. The lengthy time window may have confounded this high coherence.

Further, the proposition that microscopic signal aberrations in EEG may not be observed from a macroscopically normal EEG signal is worth considering [2]. The uncertainty of empirical, visual evaluation of EEG to localise the seizure onset zone has been shown [45]. Thus in the current work, the true onset zone for some individuals may not be observable on the raw EEG, yet may have been captured in the processed EEG coherence data. However, such postulations must be verified using high-definition EEG or intracranial/stereo EEG. Alternatively, since we combined microscopic MRI and EEG data, it is also possible that our model captured the genuine seizure onset zone. Supposing the seizure began in a different region, it could have fused in millisecond time with other active regions and thus visually revealed in a different region on the raw EEG. Such an explanation is conceivable for individuals with FBTC or multi-onset seizures and is a topic for future investigation.

Lastly, it is feasible that the structural topology has a diminished relationship with the functional activity in some individuals. Numerous mechanisms mould the seizure propagation pathways, and within-patient variance has been shown [46]. Lateralisation may be inextricably linked to the seizure duration. However, based on our experimental design (i.e. time window selection) and modest sample size, it was not plausible to extrapolate any biophysical mechanisms that may be in force. The individuals in the third group may have less well-defined circuits, or a different epileptic pattern, i.e. multi-onset or deep structural connections or functional activity, that was not captured in the connectome reconstruction or on the scalp EEG. These concepts are the subject of ongoing work to further elaborate on this feasibility study and better explain the individual differences in the outcomes. Although our methodology may explain some of the findings, potential limitations were inevitable. The generalisation of the results is limited by the small sample size, endorsing the need to extend this work to a larger cohort. The placement of the electrodes in the electrode warp and automated mapping cannot be deemed identical to the original physical placement of the electrodes during the EEG recording. The electrode warp placements represented the expected actual electrode placement during the recording. This work highlights the possible variation inherent in demystifying the electrical signals captured on scalp EEG. There is an intrinsic between-patient variance in cortex and scalp morphology and thickness. The nature of the clinical procedure introduces further variance through the measurement estimation of various technicians during the application and re-application of electrodes.

The Curry sensor coherence algorithm is confined to broadband frequencies and does not compute coherence from narrow-band frequencies. Further, the Curry algorithm does not consider the spatial, topographical, morphological or biophysical implications of the scalp signal; it is calculated purely on the raw EEG wave. Therefore, our inverse square mapping method was constrained in accounting for the spatial and biophysical properties of the scalp signal measurements. Lastly, lack of control data restricts the distinction between the structure-function coupling in seizures and normal coupling in resting state or non-ictal periods.

Indeed, linking brain structure and function remains an imperfect science, confounded by individual differences in structure-function coupling [47]. Therefore extending this study to a larger cohort, with the addition of control data, is the subject of ongoing work in our lab. The inclusion of pre-ictal and cross-hemisphere connectivity data will enable further comparison and quantification of the active electrodes across varying brain states. Additionally, using coherence matrices derived from open source software could provide a point of comparison to Curry’s sensor coherence maps.

Despite these restrictions, our work provides evidence that dMRI is a promising additional tool to classify patients for further investigation or surgery candidature. Our model may be practical in identifying the most active locations for sub-scalp electrodes and the patients who could benefit from ultra long-term monitoring. We show that all regions of high connectivity are not necessarily the best place for sub-scalp electrodes. We present a feasible method to distinguish patients, and patient-specific brain regions, that may be candidates for sub-scalp electrodes. With further refinement, our method could be utilised in identifying the optimal position for sub-scalp electrode placement, removing the need for more invasive EEG methods.

## 5. Conclusions

In conclusion, this study utilised a model to spatially map scalp electrodes to the nearest brain region and compare the structural and functional connectivity in nine patients with focal epilepsy. We showed that not all highly connected structural regions result in highly connected scalp EEG in the same region. Our findings suggest that seizures may follow strong connections intermittently and might only do so in well-lateralised patients and not for every seizure. Less well-lateralised patients displayed some high structure-function coupling in the ipsilateral hemisphere, but this was inconsistent. Our findings contribute to the evidence supporting the use of dMRI in clinical practice, which can guide patient-specific electrode placement and enhance the detection of the seizure onset zone. Future work will include comparisons with open source software and the addition of interictal and control data.

## Data Availability

The datasets generated and/or analysed during the current study are not publicly available because they are RPAH patients and can only be accessed by authorised individuals named on the approved ethics. However, de-identified, processed data can be made available upon request to the corresponding author, and subject to approval from the governing ethics entities at the RPAH and University of Sydney.

## Author Contributions

Conceptualisation, C.M., C.W., A.N.; methodology, C.M., C.W., A.N.; clinical data acquisition, C.M., A.N.; data curation, C.M., A.D.; diffusion imaging pipeline, implementation and analysis, C.M., A.D., C.W.; statistical analysis, C.M., C.W.; manuscript writing, C.M.; manuscript revision and editing C.M., A.D., M.B., O.K., C.W., A.N.; clinical advisory and results interpretation, A.N.. All authors have read and agreed to the published version of the manuscript.

## Institutional Review Board Statement

All research and methods were performed in accordance with the Declaration of Helsinki, and the relevant guidelines and regulations prescribed by the RPAH-LHD and ethics committees. The study was approved by the Ethics Committee from the RPAH Local Health District (RPAH-LHD). The protocol number and ethics approval ID for the MRI data are X14-0347 and HREC/14/RPAH/467. The protocol number and ethics approval ID for the EEG data are X19-0323 and 2019/ETH11868.

## Informed Consent Statement

The requirement for informed consent was waived in the approved ethics for the EEG data (protocol number and approval ID are X19-0323 and 2019/ETH11868) since only de-identified EEG data was acquired. Written informed consent was obtained from all participants who attended the Brain and Mind Centre for an MRI scan, as per the approved MRI ethics (protocol number and approval ID are X14-0347 and HREC/14/RPAH/467).

## Data Availability Statement

The datasets generated and/or analysed during the current study are not publicly available because they are RPAH patients and can only be accessed by authorised individuals named on the approved ethics. However, de-identified, processed data can be made available upon request to the corresponding author, and subject to approval from the governing ethics entities at the RPAH and The University of Sydney.

## Acknowledgments

The authors acknowledge all staff at the Comprehensive Epilepsy Centre at the RPAH, particularly Mrs Maricar Senturias (RN/ACNC Epilepsy), who assisted with patient recruitment. The authors acknowledge the radiology staff at i-MED Radiology for their assistance in obtaining the MRI data. The authors acknowledge the research funding support from UCB Australia Pty Ltd. CM acknowledges scholarship support from the Nerve Research Foundation, University of Sydney. AD acknowledges funding from St. Vincent’s Hospital. OK acknowledges the partial support provided by The University of Sydney through a SOAR Fellowship and Microsoft’s partial support through a Microsoft AI for Accessibility grant. CW acknowledges research funding from the Nerve Research Foundation, University of Sydney.

## Appendix A. Supplementary Figure A1

**Figure A1.**
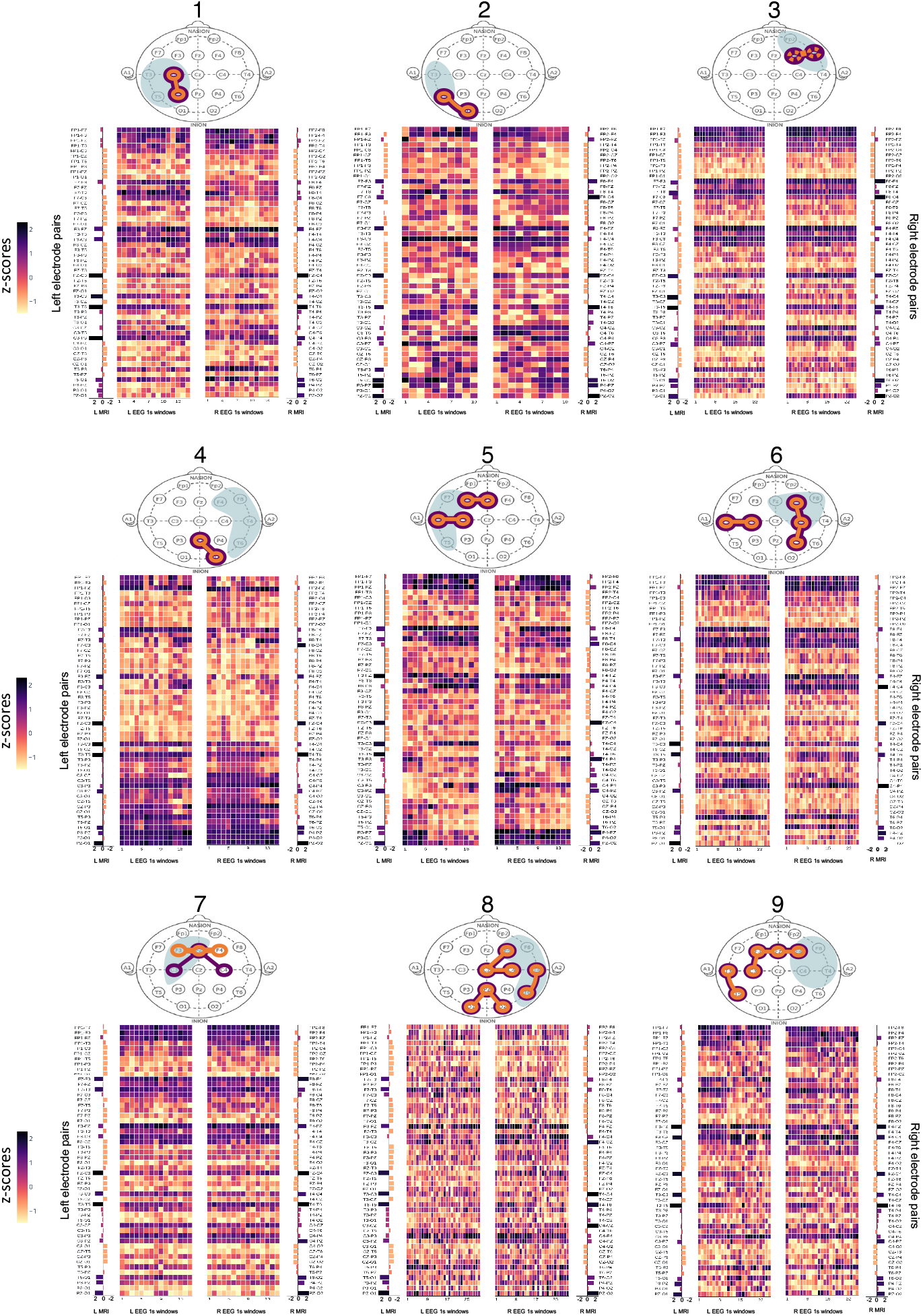
Heatmaps of z-scores from MRI and EEG connectomes. L: Left, R: Right. Patients are numbered 1-9 above each “head” and their respective connectomes are shown below the “head” map in the same grouping as Figure 2 in the main text. The “EEG 1s windows” depict the left and right side of each EEG connectome, split into 1 second windows. The structural (MRI) connectomes are split into left and right sides (“L MRI” and “R MRI”) and positioned alongside the matching EEG connectome side (i.e. left MRI next to left EEG connectome). The left and right electrode pairs are listed next to the respective side of the connectome.

